# Evaluation of statistical models of carriage to predict the impact of the 10-valent pneumococcal conjugate vaccine on invasive pneumococcal disease in Nigeria

**DOI:** 10.1101/2024.03.02.24303644

**Authors:** Aishatu L Adamu, John. Ojal, Caroline Mburu, Katherine E. Gallagher, Stefan Flasche, Kofo Odeyemi, Christy A.N. Okoromah, Isa S. Abubakar, Musa M. Bello, Victor Inem, Angela Karani, Boniface Karia, Donald Akech, Ifedayo M.O. Adetifa, J. Anthony G Scott

## Abstract

**Background:** A substantial fraction of the population-level impact of Pneumococcal Conjugate Vaccines (PCVs) on Invasive Pneumococcal Disease (IPD) is mediated through indirect effects, i.e., their capacity to protect against carriage acquisition of vaccine serotypes (VTs) among vaccinees, thereby proportionately reducing transmission and indirectly averting invasive disease in the whole population. Therefore, by relying on the consequent near elimination of VT carriage, early carriage-based models successfully captured the impact of seven-valent PCV (PCV7) in high-income settings. We sought to determine the applicability of three published statistical carriage-based models for the evaluation of PCV10 impact in Nigeria, where carriage prevalence data are available from urban and rural sites.

**Methods:** We applied external data, with assumptions, to empirical carriage prevalence data to predict IPD incidence rate ratios (IRRs). The models assume PCV has no effect on serotype invasiveness among carriers because VT carriage is eliminated. Model 1 uses estimates of relative proportions of pre-PCV VT-IPD to predict IRRs. Model 2 uses pre-PCV serotype IPD incidence, while Model 3 uses measures of serotype invasiveness, the case-carrier ratio (CCR).

**Results:** Model 1 estimates the largest PCV10 impact on overall IPD (IRR:0.38 and 0.50) in the urban and rural sites, respectively. Whereas estimates from Model 2 (IRR:0.69 and 0.78) and Model 3 (IRR:0.63 and 0.70) were more conservative.

**Conclusions:** VT carriage was not eliminated in our setting, so Model 1 estimates the hypothetical maximum impact. Relying entirely on indirect effects, Models 2 and 3 represent the minimum impact of PCV. Predictions would be more accurate if they accounted for direct effects among vaccinated VT carriers. The study illustrates the importance of capturing vaccination data on individuals sampled in carriage prevalence surveys designed to estimate IPD burden at population level.

## Introduction

Models have traditionally been used to simplify the complex relationships between host and agent in infectious disease dynamics to better understand disease burden and pathogen transmission and to predict the potential impact of interventions such as vaccines.(1) In pneumococcal disease epidemiology, models have been used across different settings to assess vaccine impact (2–5), predict the potential impact of vaccination (6,7), and guide decisions on vaccine schedules (8). The pneumococcal conjugate vaccination (PCV) protects against both pneumococcal carriage acquisition and invasion among carriers. By reducing transmission, the PCV programme has the potential for substantial indirect herd effects. Ideally, PCV impact is best demonstrated via disease endpoints measured from population-linked invasive pneumococcal disease (IPD) surveillance systems. Disease surveillance systems are, however, expensive and technically challenging to establish and sustain.(9) Therefore, pneumococcal disease surveillance is rare in low- and middle-income countries (LMICs).(10)

Models that extrapolate the impact of PCV on carriage to the impact on pneumococcal disease have been developed and validated as alternatives to disease surveillance data. These include models that incorporate complex pneumococcal transmission dynamics(2,4,7,11), those that rely on the serotype distribution and changes in carriage prevalence (12,13), and those that utilise serotype-specific carriage invasiveness.(14– 17) Dynamic models allow for the incorporation of direct and indirect effects, but their computational complexity makes them slow to develop and less widely applicable.

In the seven-valent PCV (PCV7) era, epidemiologists developed statistical carriage-based models that captured indirect vaccine effects to predict vaccine impact on IPD in high-income countries (HICs), where disease data could be used to evaluate the models.(12,14,16) In this paper, we compare the assumptions and outputs of three of these models and assess their applicability in evaluating the impact of PCVs on IPD in a low-income setting where IPD surveillance was absent.

## Methods

### Model 1-Flasche Model

Flasche *et al*. (16) proposed a model to predict the impact of PCV on total IPD incidence. This simplified version of the more complex SIS-type dynamic transmission model uses the relative prevalence of VT and NVT serotypes among carriage and disease isolates in the pre-vaccine era.(16)

In the absence of vaccination, the incidence of IPD is expressed as a carriage rate per person-time and the average risk that a carriage episode results in invasive disease, i.e., invasive capacity (IC) or case-carrier ratio (CCR), and this can be stratified into vaccine serotypes (VT) and non-vaccine serotypes (NVTs).

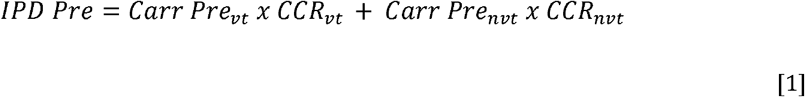

In the post-PCV period, IPD incidence can be estimated as follows:

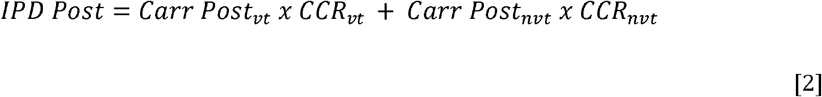

The model 1 makes three assumptions (see Table 1): (i) VT are eliminated, eventually, in the post-PCV period (*Carr post*_*vt*_ = 0), (ii) a proportion, λ, of the VT carriage will be replaced by NVT carriage (*Carr pos*_*nvt*_ = λ*Carr pre* _*vt*_ + *Carr pre*_*nvt*_), and (iii) the invasive capacity or CCR of NVT pneumococci will remain unchanged after vaccine Introduction 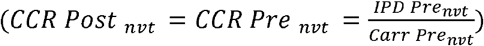. IPD incidence post-PCV can, therefore, be reformulated as follows:

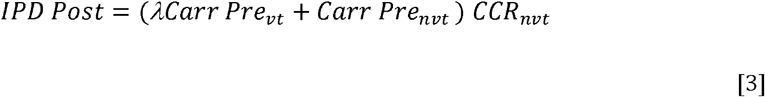

PCV impact (Incidence rate ratio [IRR]):

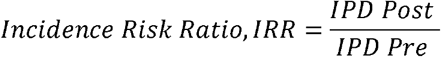

Which simplifies to (see Supplement: Appendix 1 for details):

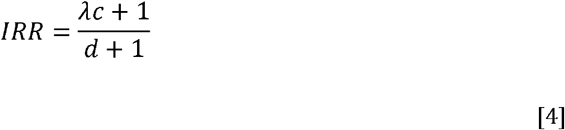

Where:

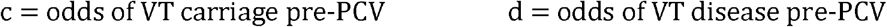

The model was validated in nine settings and demonstrated a good fit. Its predictions were robust to the introduction schedule and a wide range of PCV7 uptake levels. It was timed to three years after the introduction of PCV7 and was recently applied to estimate the global effectiveness of higher-valent PCVs.(18) The assumptions for Model 1 were valid for PCV7 introduction because it was applied predominantly to HICs where the force of infection was low, vaccine uptake was high, and introduction resulted in the rapid elimination of VT carriage and near complete replacement by non-PCV7 serotypes.

**Table 1:**
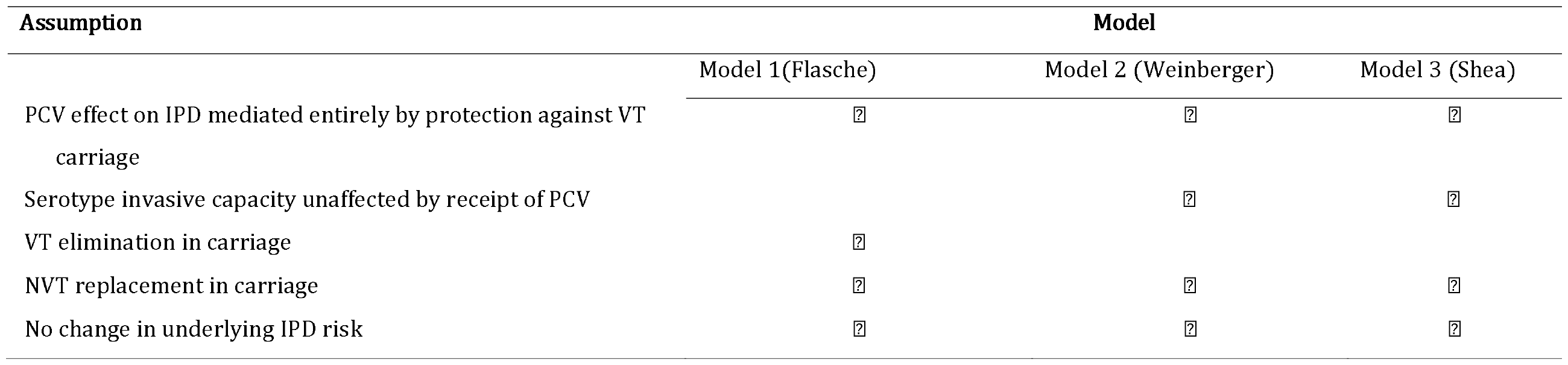
Summary of model assumptions.

| Assumption | Model |  |  |
| --- | --- | --- | --- |
|  | Model 1(Flasche) | Model 2 (Weinberger) | Model 3 (Shea) |
| PCV effect on IPD mediated entirely by protection against VT carriage | ? | ? | ? |
| Serotype invasive capacity unaffected by receipt of PCV |  | ? | ? |
| VT elimination in carriage | ? |  |  |
| NVT replacement in carriage | ? | ? | ? |
| No change in underlying IPD risk | ? | ? | ? |

### Model 2 – Weinberger Model

Weinberger *et al*. (12) proposed a model to estimate the relative changes in IPD incidence as a function of the serotype-specific pre-PCV IPD incidence and changes in carriage prevalence of the serotype. The model was initially validated using IPD incidence and carriage prevalence data from different populations (the UK, the Netherlands, USA – including Native American populations) and South Africa.(3,12)

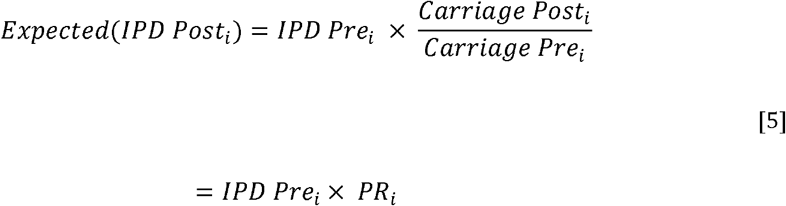

Where:

i represents individual serotypes

The key assumptions (see Table 1) of Model 2 are: (i) vaccine effectiveness against IPD is wholly accounted for by protection against carriage; (ii) there is a constant relationship between carriage and invasion, which is not affected by the vaccination status of carriers in the post-PCV era; and (ii) the underlying population IPD risk remains constant.

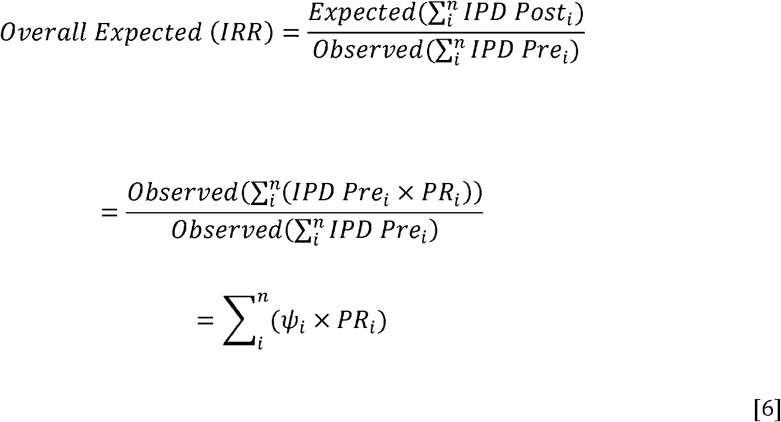

Where:

*ψ* _*i*_ = proportion of IPD in the pre-vaccine era attributable to the *i*^th^ serotype group.

The model considers changes in the prevalence of carriage in *i* strata. Applying the model where each serotype represents a single stratum did not produce optimal results when validated.(12) However, by grouping pneumococcal serotypes broadly into VT, high-incidence NVT, and low-incidence NVT, the model predicted (within 95% predictive interval) overall changes in IPD incidence as well as changes for the serotype groups.

As this model relies on carriage prevalence ratios, it assumes that all of the PCV impact on VT-IPD is mediated through reduction of VT carriage acquisition and that there is no additional direct effect of PCV on reducing VT invasive capacity. Therefore, it only accounts for vaccine protection against carriage and disregards any subsequent protection against invasion among VT carriers. Since the protection against carriage leads to a substantial indirect vaccine impact by reducing VT acquisition and VT exposure through reduced transmission, this assumption will be valid where VT carriage is eliminated. This is because eliminating VT transmission will nullify the benefit of any impact on invasion given carriage.

The assumption that the invasive capacity of each serotype remains constant after PCV introduction is justified by evidence that the invasiveness of serotypes is an intrinsic property independent of time and geography.(19,20) The assumption that vaccination does not affect invasiveness can be acceptable to some extent. We accept that PCV does not affect: (i) carriage of NVTs; (ii) invasiveness of NVTs; (iii) invasiveness of VTs among NVT carriers; and (iv) invasiveness of VTs or NVTs among non-carriers. If VT carriage is almost eliminated, the impact of PCV on the invasiveness of VT among vaccinated VT carriers will be negligible, so we can reasonably accept this assumption.

The second assumption of unchanging population susceptibility to IPD is also reasonable in HICs and when considering relatively short prediction periods that are practical for vaccine assessment (5 years), although this can vary by setting. Besides, changes in population susceptibility are likely to affect the VT and NVT disease risk to the same extent.

### Model 3 –Shea Model

Shea et al. (17) validated a third model which uses serotype-specific carriage prevalence and invasive capacity to estimate the incidence of IPD.

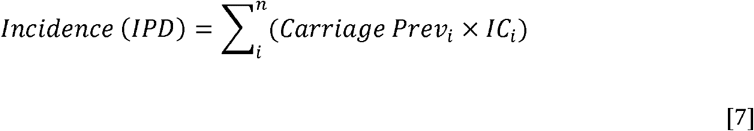

Where carriage is the instantaneous ‘prevalence of carriage’, and the IC is the ‘Invasive Capacity’ of carriage. The incidence rate ratio can be estimated if there are data on carriage prevalence in the post- and pre-vaccine era.

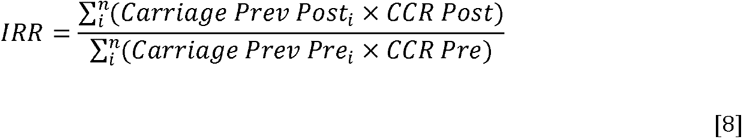

The measure of invasive capacity here is the Case-Carrier Ratio (CCR), which can be estimated from two epidemiological observations in the same population:

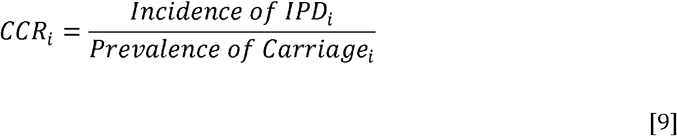

Multiplying the carriage prevalence by the CCR would give an estimate of the serotype-specific IPD incidence. When applying this model to the Nigerian settings, we have assumed that CCRs are intrinsic serotype-specific properties, which vary little with setting. Therefore, CCR values calculated from other populations can be applied to observed Nigerian carriage prevalence to estimate IPD incidence and vaccine impact. This concept has been used to predict vaccine impact on acute otitis media in the US by applying CCRs calculated from an Israeli population to US carriage prevalence data.(17)

The model also assumes that the CCR is constant in the pre- and post-PCV eras; as in model 2, the applicability of a model requiring this assumption depends on the assumption that VT carriage will largely be eliminated by the vaccine programme, as it is otherwise likely that individual vaccinees will experience a significant reduction in the risk of invasion when they become VT carriers.

We grouped serotypes into VT, high-incidence NVT, and low-incidence NVT to allow for comparison with Model 3 output,

### Relationship between Models

The *Flasche, Shea*, and Weinberger models share the assumption of independence of CCR from vaccine receipt but approach the subject from different perspectives. Drawing on different types of external data, the *Shea* and *Weinberger* models are mathematically equivalent (see Supplement: Appendix 1).

Assuming VT elimination in carriage will make all three models similar. What will differ will be the data needs for each. Model 3 requires pre and post-PCV carriage to infer disease based on external CCRs (i.e., a setting without IPD surveillance but with carriage surveys). Model 1 requires pre-PCV carriage and IPD to predict potential PCV impact on ID. Model 2 requires similar data input as Model 1 (pre-PCV disease and carriage data) but also post-PCV carriage data; making it the most data-hungry formulation and likely the most accurate if, indeed, all these data are available. The challenge for Model 2 will be the likelihood of the availability of this level of pre-PCV data but no post-PCV IPD, questioning its usefulness in reality. Another difference is that Models 2 and 3 can be formulated as either serotype-specific or serotype-group-specific, while Model 1 only uses serotype groups. For the other models, grouping serotypes is more pragmatic otherwise, important disease-causing types can be missed in carriage.

## Data sources

### Serotype-specific carriage

To estimate serotype-specific carriage prevalence in the pre-vaccine period, we used data from baseline carriage surveys conducted 4-5 months after PCV introduction in an urban and a rural site in Nigeria.(21) As PCV10 (GSK) was introduced without a catch-up campaign and as uptake was relatively modest, we estimated only a small percentage (7-15%) of children aged <5 years were vaccinated at the time of the baseline surveys, and therefore this could represent pre-vaccine epidemiology.(22) In the post-vaccine period, we conducted four (2017-2020) annual carriage surveys in the rural site and three (2018-2020) in the urban site.(22) These annual surveys used independent age-stratified random population samples and standard WHO-recommended techniques for nasopharyngeal swabbing, transport, storage and culture. Field and laboratory techniques, including the season of swabbing, were consistent across surveys.

We categorised serotypes 1, 4, 5, 6B, 7F, 9V, 14, 18C, 19F and 23F as PCV10 vaccine serotypes (VTs) and all other serotypes as non-vaccine serotypes (NVTs). In addition, we estimated serotype prevalence and proportions of VT and NVT carriage for children aged <5 years separately for the baseline and final (post-PCV) surveys in the two sites (rural and urban).

Annual pneumococcal VT and NVT carriage prevalence among children aged <5 years and the proportion of children aged <5 years vaccinated with PCV10 vaccinated are shown in Table S1. Serotype-specific carriage prevalence is shown in Figure S1.

To assess the potential of cross-protection by PCV10 against serotype 6A as reported in other settings, we did a sensitivity analysis considering a scenario where we assume that serotype 6A was a VT when categorising the serotypes.

### Serotype-specific IPD

Given that there was no direct estimate of the incidence of IPD in Nigeria in the pre-vaccine era, we obtained serotype-specific estimates of baseline IPD incidence from a systematic review of the global distribution of serotypes in IPD among children <5 years (Table S2).(23) We extracted data on serotypes from the African sub-region, which included data from 22 different studies across 13 countries spanning 1980 to 2000. Eight of the studies came from three West African countries (Burkina Faso, Mali and The Gambia), representing ∼11% of all isolates from SSA. However, 74% of the isolates from sSA were derived from South Africa (Table S3).(23)

### Measures of serotype-specific invasive capacity (case-carrier ratios, CCR)

We adopted serotype-specific CCRs (Table S4) from a meta-analysis of the ratio of IPD incidence to carriage prevalence estimated from 20 systematic and paired serotype data on asymptomatic carriage prevalence and disease samples of carriage in children.(24) A majority (15/20) of the paired studies were from countries in North America and Western Europe, and only 4/20 were from LMICs (Venezuela, Papua New Guinea and Morocco). Overall, 12/20 studies had samples from children that were strictly aged <5 years, while 8/20 studies included older (<6 years, <7 years and <18 years) children. Studies also covered the pre-(11/20) and post-PCV (9/20) periods. A summary of the number of serotypes isolated, total isolates for carriage and IPD, carriage samples and IPD surveillance population by study are shown in Table S5.

#### Estimation of Uncertainty levels of predicted IRR and IPD incidence

We calculated the uncertainty in the IRR estimates using bootstrapping and estimated the lower and upper bounds of the 95% predictive interval as 2.5% and 97.5% of 10,000 bootstrap samples for Model 1 and 1,000 for Model 2. We calculated the 95% confidence limits of the IRRs from Model 3 by adding the standard errors of the CCR and carriage prevalence using the delta method, which allows the calculation of variances of log-transformed variables.(17,25,26) Details are included in the Supplement Appendix 2.

## Results

Model 1 estimated that overall IPD incidence has declined by 50% over five years in the rural site and by 62% over four years in the urban site (Tables 2 and S6).

**Table 2:** Comparison of output from the three models showing estimated incidence rate ratios (IRRs) for impact of PCV10 on invasive pneumococcal disease in rural and urban sites in Nigeria.

|  | IRR (95% CI) |  |  |  |  |  |  |  |
| --- | --- | --- | --- | --- | --- | --- | --- | --- |
|  | VT |  | High-incidence NVT |  | Low-incidence NVT |  | Overall |  |
| Kumbotso (rural) |  |  |  |  |  |  |  |  |
| Model 1 | N/A | N/A | N/A | N/A | N/A | N/A | 0.50 | 0.47-0.54 |
| Model 2 | 0.51 | 0.35-0.72 | 0.96 | 0.65-1.42 | 1.57 | 1.23-1.97 | 0.78 | 0.65-0.95 |
| Model 3 | 0.52 | 0.44-0.61 | 0.90 | 0.73-1.10 | 1.59 | 1.26-2.00 | 0.70 | 0.67-0.73 |
| Pakoto (urban) |  |  |  |  |  |  |  |  |
| Model 1 | N/A | N/A | N/A | N/A | N/A | N/A | 0.38 | 0.36-0.39 |
| Model 2 | 0.43 | 0.31-0.58 | 1.41 | 0.86-2.30 | 1.60 | 1.38-1.85 | 0.69 | 0.58-0.83 |
| Model 3 | 0.32 | 0.26-0.40 | 0.88 | 0.70-1.13 | 1.09 | 0.88-1.40 | 0.63 | 0.60-0.67 |

Model 2 estimated that the incidence of VT-IPD declined significantly by 49% in the rural site and by 57% in the urban site (Table 2). The model also estimated a significant decline in overall IPD incidence by 22% and 31% in the two sites; IPD caused by low-incidence NVT was estimated to have increased by 57% and 60%, respectively.

Model 3 outputs estimated that overall IPD has declined by 30% in the rural site and 37% in the urban site (Table 2, Figure 1). The model also estimated a 48% and 68% decline in VT-IPD in the urban and rural sites, respectively. The model only estimated a significant increase for low-incidence NVT-IPD in the rural site.

**Figure 1:**
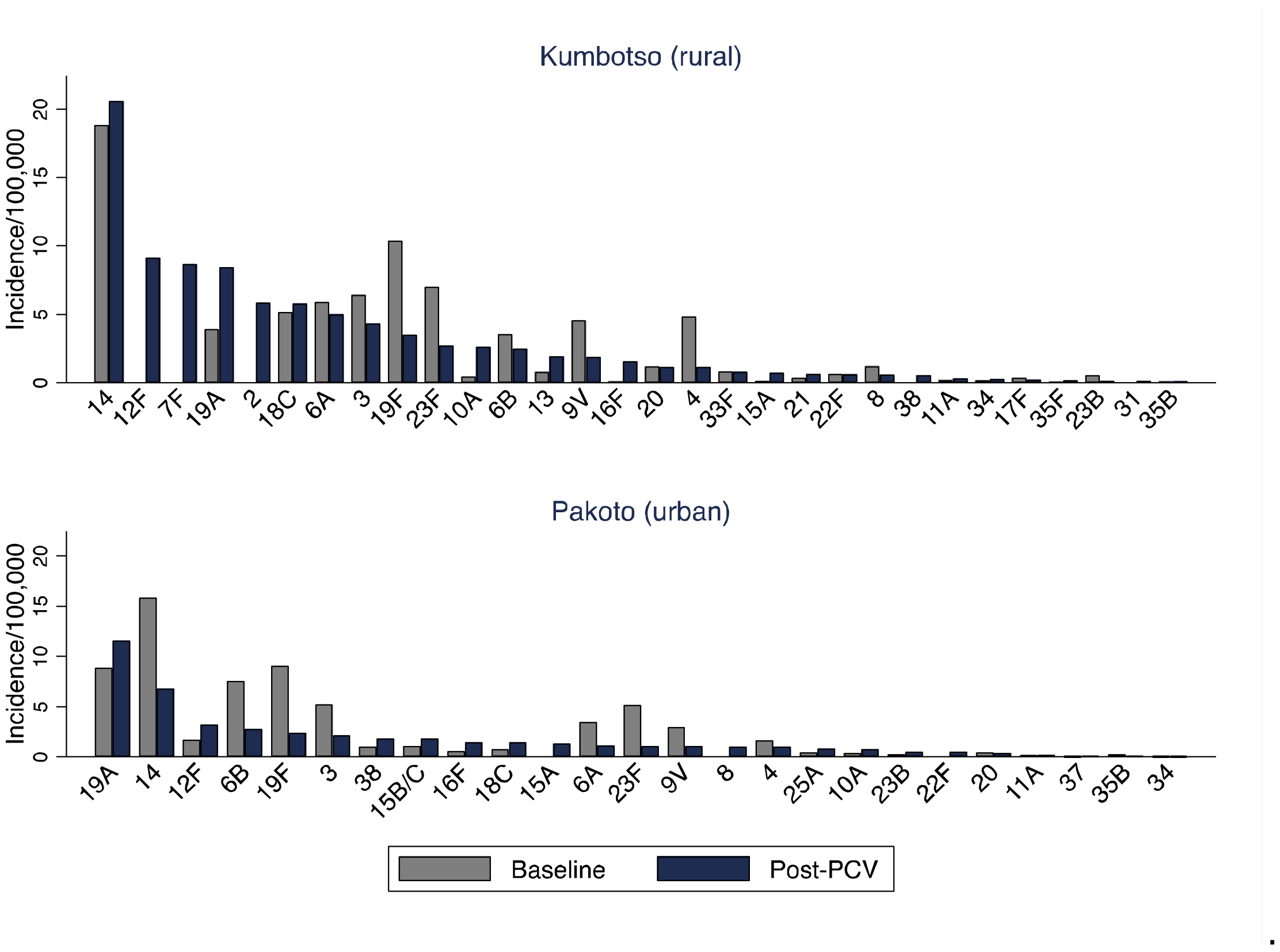
Side-by-side comparison of predicted serotype-specific incidence rates for the baseline and post-PCV periods in the rural (top) and urban (bottom) sites from Model 3. Serotypes are arranged in descending order of incidence rates in the post-PCV period.

For serotype-specific analyses from Model 3, PCV10 (and PCV10-related) serotypes dominated IPD at baseline. NVT serotypes increased in dominance post-PCV, but some VTs persisted in the predictions five years after PCV introduction (Figure 1). The serotypes predicted to cause most IPD post-PCV were 14, 12F, 7F, 19A, 2, 18C and 6A in the rural site, and 19A, 14, 12F, 6B, 19F and 3 in the urban site.

Assuming cross-protection against 6A led to larger estimates of impact in both sites for Model 1 and in the urban site for Model 2 (Table 3). Model 3 did not show any evidence of cross-protection in both sites.

**Table 3:** Sensitivity analyses exploring whether PCV10 provided cross-protection against serotype 6A*.

|  | IRR (95% CI) with Cross-protection against 6A* |  |  |  |  |  |  |  |
| --- | --- | --- | --- | --- | --- | --- | --- | --- |
|  | VT |  | High-incidence NVT |  | Low- incidence NVT |  | Overall |  |
| Kumbotso (rural) |  |  |  |  |  |  |  |  |
| Model 1 | N/A | N/A | N/A | N/A | N/A | N/A | 0.44 | 0.40-0.50 |
| Model 2 | 0.59 | 0.43-0.73 | 1.33 | 0.69-2.71 | 1.55 | 1.22-1.96 | 0.84 | 0.67-1.06 |
| Model 3 | 0.60 | 0.52-0.69 | 0.96 | 0.72-1.27 | 1.59 | 1.27-2.00 | 0.70 | 0.67-0.73 |
| Pakoto (urban) |  |  |  |  |  |  |  |  |
| Model 1 | N/A | N/A | N/A | N/A | N/A | N/A | 0.26 | 0.25-0.28 |
| Model 2 | 0.35 | 0.27-0.45 | 1.41 | 1.02-1.94 | 1.61 | 1.45-1.78 | 0.65 | 0.58-0.74 |
| Model 3 | 0.33 | 0.30-0.39 | 1.27 | 0.95-1.69 | 1.09 | 0.86-1.40 | 0.63 | 0.60-0.67 |

\* Serotype 6A was categorised as a vaccine serotype (VT)

Model 1 can only estimate overall IPD impact, and it predicted the largest relative decline in IPD incidence compared to Model 2 or Model 3. In addition to the impact on overall IPD, Models 2 and 3 also predicted the impact on VT- and NVT-IPD. The predictions from these two latter models were similar, particularly for VT and NVT-IPD in the rural site.

## Discussion

In this paper, we used three previously validated statistical carriage prevalence-based models to estimate the impact of PCV introduction on IPD in Nigeria, where IPD surveillance is lacking. These models are based on the premise that carriage is a prerequisite for invasive disease and estimate vaccine effects mediated by protection against carriage, i.e., ignore any additional direct effects of PCV on invasion. Model 1 estimated a relative decline in overall IPD incidence of 50% in the rural site and 62% in the urban site. Models 2 and 3 incorporate post-PCV VT carriage prevalence, and their predictions were substantially lower, at 22-30% and 31-37% in the rural and urban sites, respectively.

For Model 1, the assumption of independence of invasiveness from vaccine receipt does not matter eventually because post-PCV, the assumption is that VT carriage is eliminated. In our setting, this key assumption was not met. VT carriage was not eliminated; indeed, VT carriage prevalence at the end of the introduction period was 22% in the rural area and 12% in urban area.(22) In HICs, VT carriage elimination was relatively rapid and complete after PCV7 introduction (27,28) supporting the use of this simple model. By contrast, PCV13 has not eliminated the extra-non-PCV7 serotypes in HICs.(29–31). Neither PCV10 nor PCV13 has interrupted VT transmission in lower-income settings. In Mozambique, where 54% of children aged <5 years had received three doses of PCV10 three years after its introduction, VT carriage prevalence was 15-18%.(32) Even where vaccine uptake was high (>90%) or catch-up provided, VT carriage prevalence was 11% in the Gambia (33), 18% in Malawi (34) and 9% in Kenya (35) five to seven years post-PCV introduction. Thus, by overlooking this residual VT carriage and the potential disease it causes, this model inevitably underestimates the incidence of IPD post-PCV and overestimates the impact. We could interpret the model predictions as the potential impact that would have accrued if VT carriage had been eliminated in Nigeria.

Model 2 estimated a significant reduction in overall IPD of 22% and 31% in the two sites, mostly due to a reduction in VT-IPD incidence. The model does not incorporate direct protection brought about among vaccinees against invasive disease among VT carriers. Thus, its predictions for impact on VT-IPD are probably underestimated. In South Africa, the model accurately predicted impact among unvaccinated children and adults, among whom indirect effects would drive impact.(3) In contrast, among vaccinated or vaccine-eligible children in South Africa and Kenya, the model underestimated vaccine impact, indicating the impact of the model’s disregard of direct effects on invasiveness among vaccinees.(3,36)

Model 3 estimated a reduction in overall IPD of 30% and 37% in the two sites as a function of observed carriage prevalence in Nigeria and estimates of CCRs from 20 settings, none of which were in sub-Saharan Africa. This approach assumes that CCR is an intrinsic serotype characteristic (19,20), and therefore, values estimated from settings where such are available should be applicable anywhere. We have shown (Appendix 1, Supplement) how Model 3 mathematically reduces to Model 2 but uses CCRs as its input instead of prior IPD serotype distribution to interpret changes in the prevalence of carriage pre/post-vaccine. Using this approach, we predicted a significant relative decline in overall and VT-IPD incidence at nearly comparable levels to Model 2, suggesting that the different reference data selected (CCRs versus prior IPD serotype patterns) are epidemiologically comparable.

PCV protects vaccinees against VT disease via two distinct routes, i.e., mucosal protection against VT carriage acquisition and systemic protection against VT invasion following carriage. As with model 2, model 3 also assumes that receiving PCV does not affect serotype invasiveness (CCR). Therefore, the predictions underestimate vaccine impact on VTs. For NVT carriers, the CCR does not change because PCV is assumed to have no effect on the invasive capacity of NVTs. For VTs, the prediction should take account of vaccination status. Among unvaccinated VT carriers, the CCR will remain constant in the pre- and post-vaccine era. Among vaccinated VT carriers, however, the CCR is likely to be lower because vaccine-induced systemic immunity means that were the child to become a carrier, the risk of invasion will be reduced.

For Model 3 to incorporate the additional vaccine protection against invasion among vaccinees, we suggest the following adjustments to Equation 8 (see Supplement Appendix 1 for derivation):

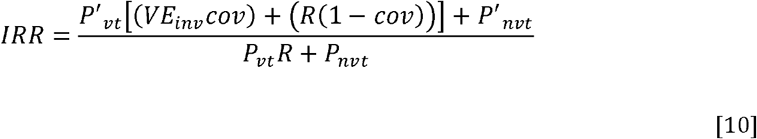

P = prevalence in the pre-vaccine era (either VT or NVT)

P’ = prevalence in the post-vaccine era (either VT or NVT)

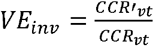, the vaccine effectiveness against invasion given carriage which is estimated as the ratio of CCR_vt_ by vaccine era,

cov = vaccine coverage among VT carriers

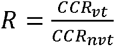, the ratio of CCRs by serotype group

Some evidence indicates that serotype-specific CCRs do not differ pre- and post-PCV (19), lending credence to the assumption for Models 2 and 3. However, for vaccinated VT carriers, VE against invasion is a function of VE against IPD and VE against carriage. Therefore, even if we accept that measured CCRs remain unchanged post-PCV, fewer PCV-driven VT carriage events at the population level will influence CCRs. Consequently, the average population CCR will be a function of the reduced VT carriage among vaccinees. In this scenario, the post-PCV CCR will still be lower than the pre-PCV CCR.

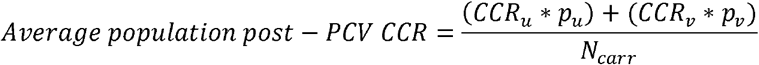

Where:

CCR_*u*_ and CCR_*v*_ = CCR among unvaccinated and vaccinated

CCR_*v*_ = CCR_*u*_ * *VE*_*inv*_,

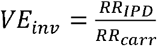,the ratio of relative reduction in IPD to relative reduction in carriage

*p*_*u*_ and *p*_*v*_= proportion of VT carriers among vaccinated and unvaccinated

*N*_*carr*_ = Number of carriers

Interpretation of pneumococcal carriage-based model predictions is subject to another constraint in addition to model-specific limitations discussed above. Our baseline carriage data were not strictly confined to the pre-PCV era because PCV10 had been introduced four to five months before the first survey; this may have led the models to underestimate the predicted impact on IPD. This effect is likely to be small because no formal catch-up was offered to older children when PCV10 was introduced, and we previously estimated that, at the time of the baseline survey, the coverage of two doses of PCV10 among all children aged <5 years was only 7% in the rural site and 15% in the urban site (see also Table S1). (22)

In conclusion, Model 1 makes a strong assumption about eliminating VT carriage, which has not been born out in the Nigerian setting examined. However, the intent of the model is not to predict exact PCV impact but rather to give an assessment on the plausible maximal impact that could be achieved if PCV programme is successful enough to largely eliminate VT carriage and transmisson. Models 2 and 3 include an analysis incorporate post-PCV VT carriage data, but they disregard direct vaccine effects against invasion by VT, which are likely to be important in contexts like Nigeria, where there is substantial residual VT carriage. Model 2 depends critically on an accurate, observed estimate of IPD incidence in the pre-vaccine period, which was unavailable for Nigeria. The accuracy of Model 3 depends on a representative estimate of the CCR, though there is little relevant data here emanating from Africa. Although these models are computationally simple and attractive for evaluating PCV impacts in sSA, these limitations undermine their general applicability. The fact that VT carriage persists in most African settings that have introduced PCV discounts the utility of Model 1 as a way to estimate current impact. The fact that there are no accurate data on the pre-PCV incidence of IPD discounts the utility of Model 2. Model 3 may be applicable if two modifications can be made; firstly, it requires credible estimates of the serotype-specific CCRs derived in populations that are representative of sSA; secondly, it requires adaptation to evaluate the direct effect of PCV among vaccinees in settings with persistent VT transmission. Unlike the deficiencies of Models 1 and 2, both of these deficiencies are amenable to further research, suggesting that Model 3 is the most propitious for evaluating PCV impact and guiding future policy in sSA using carriage studies as a proxy for complex and unaffordable IPD surveillance systems, but relies on selection of the most appropriate CCRs based on the setting, age group, and time period available.

## Data Availability

All data produced are available online at:
A. L. Adamu et al., The impact of introduction of the 10-valent pneumococcal conjugate vaccine on pneumococcal carriage in Nigeria. Nat. Commun. 14, 2666 (2023).
H. L. Johnson et al., Systematic evaluation of serotypes causing invasive pneumococcal disease among children under five: the pneumococcal global serotype project. PLoS Med. 7 (2010), doi:10.1371/journal.pmed.1000348.
A. Lochen, J. E. Truscott, N. J. Croucher, Analysing pneumococcal invasiveness using Bayesian models of pathogen progression rates. PLoS Comput. Biol. 18, e1009389 (2022).

https://journals.plos.org/ploscompbiol/article?id=10.1371/journal.pcbi.1009389

https://www.nature.com/articles/s41467-023-38277-z

## Ethics approval

This Nigeria carriage prevalence data utilised in the models is from a study that involves human participants and was approved by Aminu Kano Teaching Hospital Research Ethics Committee -NHREC/21/08/2008/AKTH/EC/2165 and Kano State Ministry of Health Research Ethics Committee - MOH/OFF/797/T.I/596, Kenya Medical Research Institute Scientific and Ethical Review Unit SERU 3350, London School of Hygiene and Tropical Medicine Observational/Interventions Research Ethics Committee - Ref 11670. Participants gave informed consent to participate in the study before taking part.

## Data availability

The authors declare that data supporting the findings were obtained from previously published work that is openly available and included within the paper and its supplementary information files (Supplementary Data).

## References

1. Grassly NC, Fraser C. Mathematical models of infectious disease transmission. Nat Rev Microbiol. 2008 Jun;6(6):477–87.

2. Melegaro A, Choi YH, George R, Edmunds WJ, Miller E, Gay NJ. Dynamic models of pneumococcal carriage and the impact of the Heptavalent Pneumococcal Conjugate Vaccine on invasive pneumococcal disease. BMC Infect Dis. 2010 Apr 8;10:90.

3. Nzenze SA, Madhi SA, Shiri T, Klugman KP, de Gouveia L, Moore DP, et al. Imputing the direct and indirect effectiveness of childhood pneumococcal conjugate vaccine against invasive pneumococcal disease by surveying temporal changes in nasopharyngeal pneumococcal colonization. Am J Epidemiol. 2017 Aug 15;186(4):435–44.

4. Ojal J, Griffiths U, Hammitt LL, Adetifa I, Akech D, Tabu C, et al. Sustaining pneumococcal vaccination after transitioning from Gavi support: a modelling and cost-effectiveness study in Kenya. Lancet Glob Health. 2019;7(5):e644–54.

5. Rinta-Kokko H, Nurhonen M, Auranen K. Impact and effectiveness of a conjugate vaccine against invasive pneumococcal disease in Finland - a modelling approach. Hum Vaccin Immunother. 2021 Jun 3;17(6):1834–43.

6. Ghia C, Wasserman M, Fletcher M, Farkouh R, Rambhad G. Modeling Possible Inclusion of Pneumococcal Conjugate Vaccine into the National Immunization Program for Infants in India. Value in Health Regional Issues. 2018 May;15:99–105.

7. Le Polain De Waroux O, Edmunds WJ, Takahashi K, Ariyoshi K, Mulholland EK, Goldblatt D, et al. Predicting the impact of pneumococcal conjugate vaccine programme options in Vietnam. Hum Vaccin Immunother. 2018 Jun 8;14(8):1939–47.

8. Choi YH, Andrews N, Miller E. Estimated impact of revising the 13-valent pneumococcal conjugate vaccine schedule from 2+1 to 1+1 in England and Wales: A modelling study. PLoS Med. 2019 Jul 3;16(7):e1002845.

9. Mackenzie GA, Plumb ID, Sambou S, Saha D, Uchendu U, Akinsola B, et al. Monitoring the introduction of pneumococcal conjugate vaccines into West Africa: design and implementation of a population-based surveillance system. PLoS Med. 2012 Jan 17;9(1):e1001161.

10. Deloria Knoll M, Bennett JC, Garcia Quesada M, Kagucia EW, Peterson ME, Feikin DR, et al. Global Landscape Review of Serotype-Specific Invasive Pneumococcal Disease Surveillance among Countries Using PCV10/13: The Pneumococcal Serotype Replacement and Distribution Estimation (PSERENADE) Project. Microorganisms. 2021 Apr 2;9(4).

11. Ojal J, Flasche S, Hammitt LL, Akech D, Kiti MC, Kamau T, et al. Sustained reduction in vaccine-type invasive pneumococcal disease despite waning effects of a catch-up campaign in Kilifi, Kenya: A mathematical model based on pre-vaccination data. Vaccine. 2017 Aug 16;35(35 Pt B):4561–8.

12. Weinberger DM, Bruden DT, Grant LR, Lipsitch M, O’Brien KL, Pelton SI, et al. Using pneumococcal carriage data to monitor postvaccination changes in invasive disease. Am J Epidemiol. 2013 Nov 1;178(9):1488–95.

13. Flasche S, Givon-Lavi N, Dagan R. Using pneumococcal carriage data to monitor postvaccination changes in the incidence of pneumococcal otitis media. Am J Epidemiol. 2016 Nov 1;184(9):652–9.

14. Weinberger DM, Harboe ZB, Flasche S, Scott JA, Lipsitch M. Prediction of serotypes causing invasive pneumococcal disease in unvaccinated and vaccinated populations. Epidemiology. 2011 Mar;22(2):199–207.

15. Weinberger DM, Grant LR, Weatherholtz RC, Warren JL, O’Brien KL, Hammitt LL. Relating pneumococcal carriage among children to disease rates among adults before and after the introduction of conjugate vaccines. Am J Epidemiol. 2016 Jun 1;183(11):1055–62.

16. Flasche S, Le Polain de Waroux O, O’Brien KL, Edmunds WJ. The serotype distribution among healthy carriers before vaccination is essential for predicting the impact of pneumococcal conjugate vaccine on invasive disease. PLoS Comput Biol. 2015 Apr 16;11(4):e1004173.

17. Shea KM, Weycker D, Stevenson AE, Strutton DR, Pelton SI. Modeling the decline in pneumococcal acute otitis media following the introduction of pneumococcal conjugate vaccines in the US. Vaccine. 2011 Oct 1;29(45):8042–8.

18. Chen C, Cervero Liceras F, Flasche S, Sidharta S, Yoong J, Sundaram N, et al. Effect and cost-effectiveness of pneumococcal conjugate vaccination: a global modelling analysis. Lancet Glob Health. 2019 Jan;7(1):e58–67.

19. Scott JR, Millar EV, Lipsitch M, Moulton LH, Weatherholtz R, Perilla MJ, et al. Impact of more than a decade of pneumococcal conjugate vaccine use on carriage and invasive potential in Native American communities. J Infect Dis. 2012 Jan 1;205(2):280–8.

20. Brueggemann AB, Peto TEA, Crook DW, Butler JC, Kristinsson KG, Spratt BG. Temporal and geographic stability of the serogroup-specific invasive disease potential of Streptococcus pneumoniae in children. J Infect Dis. 2004 Oct 1;190(7):1203–11.

21. Adetifa IMO, Adamu AL, Karani A, Waithaka M, Odeyemi KA, Okoromah CAN, et al. Nasopharyngeal Pneumococcal Carriage in Nigeria: a two-site, population-based survey. Sci Rep. 2018 Feb 22;8(1):3509.

22. Adamu AL, Ojal J, Abubakar IA, Odeyemi KA, Bello MM, Okoromah CAN, et al. The impact of introduction of the 10-valent pneumococcal conjugate vaccine on pneumococcal carriage in Nigeria. Nat Commun. 2023 May 9;14(1):2666.

23. Johnson HL, Deloria-Knoll M, Levine OS, Stoszek SK, Freimanis Hance L, Reithinger R, et al. Systematic evaluation of serotypes causing invasive pneumococcal disease among children under five: the pneumococcal global serotype project. PLoS Med. 2010 Oct 5;7(10).

24. Løchen A, Truscott JE, Croucher NJ. Analysing pneumococcal invasiveness using Bayesian models of pathogen progression rates. PLoS Comput Biol. 2022 Feb 17;18(2):e1009389.

25. Yildirim I, Hanage WP, Lipsitch M, Shea KM, Stevenson A, Finkelstein J, et al. Serotype specific invasive capacity and persistent reduction in invasive pneumococcal disease. Vaccine. 2010 Dec 16;29(2):283–8.

26. Kirkwood BR, Sterne JA. Comparing two proportions. Essential medical statistics. John Wiley & Sons; 2010. p. 148–64.

27. Flasche S, Van Hoek AJ, Sheasby E, Waight P, Andrews N, Sheppard C, et al. Effect of pneumococcal conjugate vaccination on serotype-specific carriage and invasive disease in England: a cross-sectional study. PLoS Med. 2011 Apr 5;8(4):e1001017.

28. Lee GM, Kleinman K, Pelton SI, Hanage W, Huang SS, Lakoma M, et al. Impact of 13-Valent Pneumococcal Conjugate Vaccination on Streptococcus pneumoniae Carriage in Young Children in Massachusetts. J Pediatric Infect Dis Soc. 2014 Mar;3(1):23–32.

29. Kandasamy R, Voysey M, Collins S, Berbers G, Robinson H, Noel I, et al. Persistent Circulation of Vaccine Serotypes and Serotype Replacement After 5 Years of Infant Immunization With 13-Valent Pneumococcal Conjugate Vaccine in the United Kingdom. J Infect Dis. 2020 Mar 28;221(8):1361–70.

30. Tiley KS, Ratcliffe H, Voysey M, Jefferies K, Sinclair G, Carr M, et al. Nasopharyngeal Carriage of Pneumococcus in Children in England up to 10 Years After 13-Valent Pneumococcal Conjugate Vaccine Introduction: Persistence of Serotypes 3 and 19A and Emergence of 7C. J Infect Dis. 2023 Mar 1;227(5):610–21.

31. Løvlie A, Vestrheim DF, Aaberge IS, Steens A. Changes in pneumococcal carriage prevalence and factors associated with carriage in Norwegian children, four years after introduction of PCV13. BMC Infect Dis. 2020 Jan 10;20(1):29.

32. Valenciano SJ, Moiane B, Lessa FC, Chaúque A, Massora S, Pimenta FC, et al. Effect of 10-Valent Pneumococcal Conjugate Vaccine on Streptococcus pneumoniae Nasopharyngeal Carriage Among Children Less Than 5 Years Old: 3 Years Post-10-Valent Pneumococcal Conjugate Vaccine Introduction in Mozambique. J Pediatric Infect Dis Soc. 2021 Apr 30;10(4):448–56.

33. Usuf E, Bottomley C, Bojang E, Cox I, Bojang A, Gladstone R, et al. Persistence of nasopharyngeal pneumococcal vaccine serotypes and increase of nonvaccine serotypes among vaccinated infants and their mothers 5 years after introduction of pneumococcal conjugate vaccine 13 in the gambia. Clin Infect Dis. 2019 Apr 24;68(9):1512–21.

34. Swarthout TD, Fronterre C, Lourenço J, Obolski U, Gori A, Bar-Zeev N, et al. High residual carriage of vaccine-serotype Streptococcus pneumoniae after introduction of pneumococcal conjugate vaccine in Malawi. Nat Commun. 2020 May 6;11(1):2222.

35. Hammitt LL, Etyang AO, Morpeth SC, Ojal J, Mutuku A, Mturi N, et al. Effect of ten-valent pneumococcal conjugate vaccine on invasive pneumococcal disease and nasopharyngeal carriage in Kenya: a longitudinal surveillance study. Lancet. 2019 May 25;393(10186):2146–54.

36. Adetifa IMO, Ojal J, Hammitt LI, Morpeth SC, Akech DO, Karani A, et al. Using pneumococcal carriage data to predict post-vaccination changes to invasive disease in rural Kenya - a validation exercise using Kilifi data. Glasgow, Scotland: ISPPD; 2016.

