## Supplement for "Evaluation of statistical models of carriage to predict the impact of the 10-valent pneumococcal conjugate vaccine on invasive pneumococcal disease in Nigeria"

**Appendix 1: Equations**

**Model 1**

IPD incidence in the pre and post-PCV are estimated as

$$IPD Pre={Carr Pre}_{vt} x {CCR}_{vt}+ {Carr Pre}_{nvt} x {CCR}_{nvt}$$

$$IPD Post={Carr Post}_{vt} x {CCR}_{vt}+ {Carr Post}_{nvt} x {CCR}_{nvt}$$

Assuming:

i. VT elimination, ${Carr Post}_{vt}=0$
ii. NVT replacement λ, such that ${(Carr Post}_{nvt}= \lambda{Carr Pre}_{vt}+{Carr Pre}_{nvt})$
iii. unchanged NVT invasiveness post-PCV

$${(CCR Post}_{nvt}={CCR Pre}_{nvt}=\frac{{IPD Pre}_{nvt}}{{Carr Pre}_{nvt}})$$

Then:

$$IPD Post={(\lambda Carr Pre}_{vt}+{Carr Pre}_{nvt}) {CCR}_{nvt}$$

PCV impact (Incidence rate ratio [IRR]):

$$Incidence Risk Ratio, IRR=\frac{IPD Post}{IPD Pre}$$

$$IRR=\frac{{(\lambda Carr Pre}_{vt}+{Carr Pre}_{nvt}) {CCR}_{nvt}}{{Carr Pre}_{vt} x {CCR}_{vt}+ {Carr Pre}_{nvt} x {CCR}_{nvt}}$$

IRR is then simplified to pre-PCV VT carriage and disease odds.

$$IRR=\frac{\lambda(\frac{{Carr Pre}_{vt}}{{Carr Pre}_{nvt}})+1}{(\frac{{{IPD Pre}_{vt}}}{{IPD Pre}_{nvt}})+1}$$

$$IRR=\frac{\lambda c+1}{d+1}$$

Where:

c = odds of VT carriage pre-PCV	d = odds of VT disease pre-PCV

**Relationship between Models 2 and 3**

In both cases, this is likely to be wrong, and the effect of the vaccine in preventing invasion, conditional upon carriage, is likely to be strong, particularly in the presence of residual VT carriage. Hence, both models will underestimate the total impact of the vaccine introduction.

If we use an estimate of IC from the pre-vaccine era,

$${IC}_{i}=\frac{{IPD Pre}_{i}}{{Carriage Pre}_{i}}$$

Then the *Shea* model resolves into the Weinberger model.

$$IRR=\frac{\sum_{i}^{n} ({Carriage Post}_{i}\times{IC}_{i})}{\sum_{i}^{n} ({Carriage Pre}_{i}\times{IC}_{i})}$$

$$IRR=\frac{\sum_{i}^{n} ({Carriage Post}_{i}\times{IPD Pre}_{i}/{Carriage Pre}_{i})}{\sum_{i}^{n} ({Carriage Pre}_{i}\times{IPD Pre}_{i}/{Carriage Pre}_{i}))}$$

$$IRR=\frac{\sum_{i}^{n} ({{IPD Pre}_{i}\times Carriage Post}_{i}/{Carriage Pre}_{i})}{\sum_{i}^{n} ({IPD Pre}_{i}))}$$

$$IRR=\frac{\sum_{i}^{n} ({IPD Pre}_{i}\times{PR}_{i})}{\sum_{i}^{n} ({IPD Pre}_{i})}$$

**Modification of Model 3 to account for direct vaccine effects among vaccinated VT carriers**

To estimate IPD, we would need to account for the reduced invasiveness among vaccinees who are VT carriers. Model 3 estimates IPD as a function of respective carriage prevalence and CCR in the pre and post-PCV periods, assuming CCR is constant across the periods.

$$IRR=\frac{{Carriage Prev Post}\times CCR}{{Carriage Prev Pre}\times CCR}$$

The equation above can be expanded to show how VT and NVT groups are incorporated, with the assumption that $CCR Pre=CCR Post$ for VTs and NVTs:

$$IRR=\frac{\left( {Carr Prev Post}_{vt}\times CCR {Post}_{vt} \right)+(Carr {Prev Post}_{nvt} {CCR}_{nvt})}{\left( {Carr Prev Pre}_{vt}\times CCR {Pre}_{vt} \right)+ ({Carr Prev Pre}_{nvt}\times CCR {Pre}_{nvt})}$$

We adjust the equation above to incorporate different CCRs for VT carriage and vaccine uptake:

$$IRR=\frac{cov{(Carr Prev Post}_{vt}\times{CCR Post}_{vt(v)})+\left( 1-cov \right)\left( {Carr Prev Post}_{vt}\times{CCR}_{vt\left( u \right)} \right)+(Carr {Prev Post}_{nvt} {CCR}_{nvt})}{({Carr Prev Pre}_{vt} x {CCR}_{vt\left( u \right)})+({Carr Prev Pre}_{nvt} x {CCR}_{nvt})}$$

Where:

vac cov = vaccine coverage in the target population (children <5 years) in the post-vaccine era (among the children in the carriage survey)

Carriage in the pre- and post-PCV periods can be broken down into VT and NVT carriage

CCR = case carrier ratio, which is different for VT and for NVT and also varies for VT, depending on whether the child has been vaccinated (v) or unvaccinated (u); all VT carriers in the pre-vaccine period experience the CCR for VT of unvaccinated individuals. Vaccination has no impact on the CCR for NVT

To simplify the equation, we divide the subparts of the equation by a common term, i.e., CCR_nvt_

$$IRR=\frac{cov\left( \frac{{Carr Prev Post}_{vt}\times{CCR Post}_{vt\left( v \right)}}{{CCR}_{nvt}} \right)+\left( 1-cov \right)(\frac{{Carr Prev Post}_{vt}\times{CCR}_{vt\left( u \right)}}{{CCR}_{nvt}})+\frac{Carr {Prev Post}_{nvt} {CCR}_{nvt}}{{CCR}_{nvt}}}{({\frac{{Carr Prev Pre}_{vt} \times{CCR}_{vt\left( u \right)}}{{CCR}_{nvt}} )}+(\frac{{Carr Prev Pre}_{nvt}\times{CCR}_{nvt}}{{CCR}_{nvt}})}$$

$$IRR=\frac{cov\left( {P'}_{vt}\times{\frac{{CCR^{'}}_{vt}}{{CCR}_{nvt}}} \right)+\left( 1-cov \right)( {P'}_{vt}\times{\frac{{CCR}_{vt}}{{CCR}_{nvt}}})+ {P'}_{nvt}\times{\frac{{CCR}_{nvt}}{{CCR}_{nvt}}}}{({P_{vt}\times{\frac{{CCR}_{vt}}{{CCR}_{nvt}}} )}+(P_{nvt}\times{\frac{{CCR}_{nvt}}{{CCR}_{nvt}}})}$$

$$IRR=\frac{{P'}_{vt}[\left( {VE}_{inv}\times cov \right)+R\left( 1-cov \right)]+ {P'}_{nvt}}{{P_{vt}R}+P_{nvt}}$$

Where:

P = prevalence in the pre-vaccine era (either VT or NVT)

P’ = prevalence in the post-vaccine era (either VT or NVT)

VE_inv_ = the ratio of CCR_vt_ by vaccine era, hence CCR’_vt_/CCR_vt_

cov = vaccine coverage among VT carriers

R = the ratio of CCRs, hence CCR_vt_/CCR_nvt_

Note the equation can be further expanded to include further breakdown of NVTs into high-incidence and low0incidence NVTs.

Alternatively, we could also divide the equation sub-parts by CCR_vt(u)_

$$IRR=\frac{cov\left( \frac{{Carr Prev Post}_{vt}\times{CCR}_{vt\left( v \right)}}{{CCR}_{vt\left( u \right)}} \right)+\left( 1-cov \right)\left( \frac{{Carr Prev Post}_{vt}\times{CCR}_{vt\left( u \right)}}{{CCR}_{vt\left( u \right)}} \right)+\frac{Carr {Prev Post}_{nvt} {CCR}_{nvt}}{{CCR}_{vt\left( u \right)}}}{({\frac{{Carr Prev Pre}_{vt} x {CCR}_{vt\left( u \right)}}{{CCR}_{vt\left( u \right)}} )}+(\frac{{Carr Prev Pre}_{nvt} x {CCR}_{nvt}}{{CCR}_{vt\left( u \right)}})}$$

$$IRR=\frac{cov\left( {P'}_{vt}\times{\frac{{CCR^{'}}_{vt}}{{CCR}_{vt}}} \right)+\left( 1-cov \right)( {P'}_{vt}\times{\frac{{CCR}_{vt}}{{CCR}_{vt}}})+ {P'}_{nvt}\times{\frac{{CCR}_{nvt}}{{CCR}_{vt}}}}{({P_{vt}\times{\frac{{CCR}_{vt}}{{CCR}_{vt}}} )}+(P_{nvt}\times{\frac{{CCR}_{nvt}}{{CCR}_{vt}}})}$$

$$IRR=\frac{{P'}_{vt}[\left( {VE}_{inv}\times cov \right)+\left( 1-cov \right)]+ {P'}_{nvt}R}{{P_{vt}{}}+P_{nvt}R}$$

Where:

VE = CCR’_vt_/CCR_vt_

R = CCR_nvtt_/CCR_vt_

**Appendix 2: Estimation of Uncertainty levels of estimated Incidence rate Ratios (IRRs)**

Model 1

To calculate the predicted IRRs and their corresponding uncertainties, we assumed that the proportion of VTs among both carriers and IPD isolates were samples from binomial distributions and drew 10,000 bootstrap samples. Then, we calculated the median value of these draws as the point estimate of the IRR and the 2.5% and 97.5% of the draws as lower and upper bounds of the 95% predictive interval (PI).

Model 2

First, we calculated the prevalence ratios (post-PCV divided by the baseline carriage prevalence) of individual serotypes. The baseline carriage data used were carriage surveys for 2016 (rural site) and 2017 (urban site), while the post-PCV carriage data used were carriage surveys for 2020 in both sites. For serotypes not observed in carriage in the baseline and post-PCV surveys, we did a continuity correction (adding 0.5 to the numerator and denominator as suggested by Weinberger) prior to calculating the prevalence ratios (PR).

We conducted this analysis in three strata of serotype groups; VT, high-incidence NVT and low-incidence NVT. We classified serotypes into these groups based on a meta-analysis of serotype-specific IPD incidence for Africa(1). The authors reported IPD incidence for 21 individual serotypes, including all ten vaccine serotypes in PCV10. We classified the 11 NVTs with incidence rates sufficiently high to be calculated individually as ‘high incidence NVT’. For all other serotypes, the serotype-specific incidence was too low to be estimated, and we defined these as ‘low incidence NVT’.

We calculated the incidence rate ratios (IRR) using the prevalence ratios and pre-vaccination IPD proportions for each serotype. We assessed the uncertainty inherent in the IRR via probabilistic resampling, i.e., bootstrapping. For each of the 1000 bootstrap samples, we assumed a multivariate normal distribution of the log of prevalence ratios with mean and variance estimated from a weighted regression model. For the IPD incidence, we assumed a Poisson distribution with the mean equal to the pre-PCV IPD incidence of each serotype. We estimated the average predicted post-PCV IPD incidence as a product of the drawn pre-PCV IPD and weighted PR by the serotype groups (VT/high-incidence NVT/low-incidence NVT). We calculated the IRR by dividing the predicted post-PCV IPD incidence by the drawn pre-PCV IPD incidence for the three serotype groups. We calculated the median and the 2.5% and 97.5% of the predicted IRRS (for VT and NVT) from 1,000 draws as the point estimate and 95% predictive interval, respectively.

Model 3

The 95% confidence limits of the IRRs were calculated by adding the standard errors of the IC and carriage prevalence using the delta method, which allows the calculation of variances of log-transformed variables. The variance of the natural logarithms of the ICs for each serotype are normally distributed and calculated as follows:(2,3)

$$\frac{1}{No of IPD cases of serotype i}+\frac{1-Carr prevalence of serotype i}{Carr prevalence of serotype i x No of children swabbed}$$

The standard error of carriage prevalence is calculated using the delta method, where the standard error of the natural logarithm of a proportion is calculated as follows:(4)

$$\frac{1}{Number of carriers}- \frac{1}{Number of children swabbed}$$

Where 95% confidence intervals of IPD incidence are calculated as the exponent of:

$$Log IPD incidence \pm1.96 x standard error\left( SE \right)of\log IPD incidence$$

Where:

$$SE\log IPD incidenc=SE of IC+SE of carriage prevalence$$

**Supplementary Tables and Figures**

Table S1: Annual observed uptake of three doses of PCV10 and pneumococcal carriage (overall and vaccine type, VT) prevalence in the rural and urban sites by survey year

|  | **PCV10 coverage (95% CI)** | | **Overall carriage (95% CI)** | **VT carriage (95% CI)** |
| --- | --- | --- | --- | --- |
| **Rural** | | | | |
| Year 1^1^ | | 7 (6-9) | 91 (88-94) | 42 (37-48) |
| Year 2 | | 28 (22-34) | 92 (89-96) | 30 (25-36) |
| Year 3 | | 57 (53-61) | 92 (90-95) | 25 (21-30) |
| Year 4 | | 69 (64-73) | 91 (88-94) | 21 (17-26) |
| Year 5^2^ | | 59 (51-67) | 88 (84-91) | 22 (18-27) |
| **Urban** | | | | |
| Year 1^3^ | | 15 (13-17) | 77 (72-81) | 38 (33-43) |
| Year 2 | | 61 (57-65) | 70 (65-76) | 23 (18-29) |
| Year 3 | | 75 (71-80) | 69 (63-75) | 19 (15-25) |
| Year 4^2^ | | 81 (74-86) | 53 (46-61) | 12 (8-17) |

^1^ Baseline carriage data included in the models. Survey was conducted in 2016, five months after PCV10 was introduced

^2^ Post-PCV carriage data included in the models. Survey was conducted in 2020.

^3^ Baseline carriage data included in the models. Survey was conducted in 2017, four months after PCV10 was introduced

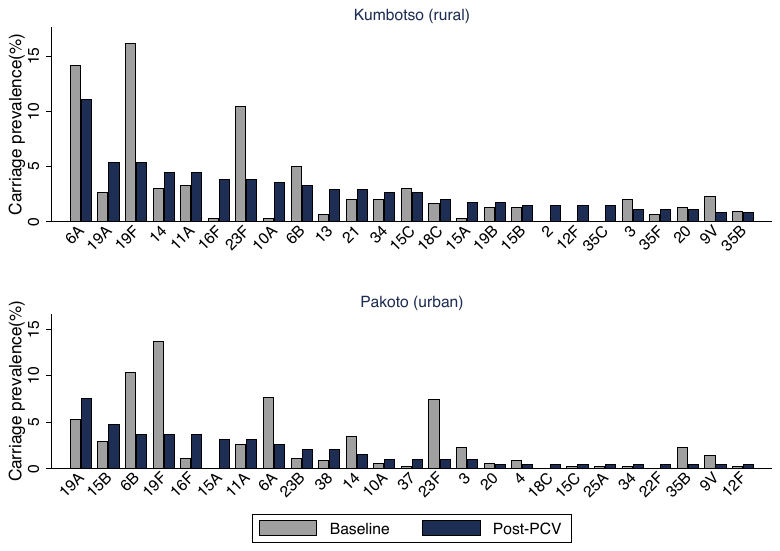

Figure S1: Serotype-specific carriage prevalence in the baseline and post-PCV periods in the rural (top) and urban (bottom) sites. Serotypes arranged in descending of prevalence levels in the post-PCV period.

Table S2: Proportions (%) and incidence rates (per 100,000) in IPD attributable to serotypes in children aged <5 years in Africa in the pre-PCV era as reported by Johnson et al.(1)

| Serotype | Proportion (%) | 95% CI | Incidence/100,000 | LB, UB |
| --- | --- | --- | --- | --- |
| 1 | 11.70% | 9.5, 13.8 | 396 | 243, 582 |
| 2 | 1.90% | 1.0, 2.8 | 65 | 25, 119 |
| 3 | 1.10% | 0.8, 1.5 | 38 | 20, 61 |
| 4 | 2.30% | 1.7, 3.0 | 79 | 43, 125 |
| 5 | 10.70% | 7.6, 13.8 | 364 | 193, 584 |
| 6A | 9.40% | 7.2, 11.5 | 317 | 182, 488 |
| 6B | 8.50% | 6.3, 10.7 | 288 | 160, 451 |
| 7F | 0.80% | 0.4, 1.3 | 28 | 10, 54 |
| 8 | 1.10% | 0.8, 1.5 | 38 | 19, 63 |
| 9A | 0.40% | 0.2, 0.7 | 15 | 6, 28 |
| 9V | 2.20% | 1.3, 3.1 | 74 | 34, 129 |
| 12A | 0.10% | 0.0, 0.1 | 2 | 0, 5 |
| 12F | 1.70% | 1.1, 2.3 | 57 | 27, 99 |
| 14 | 13.00% | 10.0, 16.0 | 441 | 254, 676 |
| 15B | 0.50% | 0.1, 0.9 | 18 | 3, 39 |
| 18C | 1.40% | 0.9, 2.0 | 48 | 22, 84 |
| 19A | 3.90% | 2.5, 5.3 | 133 | 63, 226 |
| 19F | 5.40% | 3.6, 7.1 | 182 | 92, 300 |
| 23F | 6.50% | 4.5, 8.5 | 220 | 114, 359 |
| 45 | 0.50% | 0.0, 1.0 | 17 | 0, 44 |
| 46 | 1.30% | 0.4, 2.1 | 43 | 10, 90 |
| All Others | 15.70% | 12.7, 18.6 | 532 | 322, 790 |
| TOTAL | 100.00% |  | 3,395 |  |

CI=Confidence Interval; LB=Lower bound of uncertainty estimate; UB=Upper bound of uncertainty estimate

Table S3: Countries in Africa that contributed to IPD serotype data in Table S2 and numbers of isolates contributed. (1)

|  | Country | Study years | Total No. isolates |
| --- | --- | --- | --- |
| AFRICA (N=22 studies) | |  |  |
| 1. | Algeria | 1996-2000 | 45 |
| 2. | Burkina Faso | 2002-2005 | 22 |
| 3. | Egypt | 1998-2003 | 113 |
| 4. | Ethiopia | 1993-1995 | 46 |
| 5. | Kenya | 1994-2007 | 595 |
| 6. | Kenya | 2004-2007 | 46 |
| 7. | Malawi | 1996-1998 | 122 |
| 8. | Mali | 2003-2004 | 54 |
| 9. | Mali | 2002-2007 | 570 |
| 10. | Mozambique | 2001-2007 | 259 |
| 11. | Rwanda | 1984-1990 | 130 |
| 12. | South Africa | 1989-1991 | 181 |
| 13. | South Africa | 1993-1995 | 98 |
| 14. | South Africa | 1998-2001 | 66 |
| 15. | South Africa | 2000-2006 | 8221 |
| 16. | Tanzania | 2006-2007 | 27 |
| 17. | The Gambia | 1993-1995 | 105 |
| 18. | The Gambia | 1989-1991 | 60 |
| 19. | The Gambia | 1996-2003 | 212 |
| 20. | The Gambia | 2000-2003 | 116 |
| 21. | The Gambia | 1990-1992 | 46 |
| 22. | Uganda | 2004-2007 | 47 |

Table S4: Serotype-specific case-carrier ratios estimated for children from meta-analysis reported by Lochen et al (5) and underlying isolates for carriage and IPD

| Serotype* | CCR | Carriage isolates | IPD isolates |
| --- | --- | --- | --- |
| 1 | 0.01736978 | 25 | 176 |
| 7F | 0.00958808 | 51 | 172 |
| 5 | 0.00732476 | 7 | 31 |
| 12F | 0.00605978 | 26 | 45 |
| 27 | 0.00449171 | 2 | 6 |
| 14 | 0.00426382 | 344 | 511 |
| 2 | 0.00389772 | 2 | 2 |
| 18C | 0.00273942 | 48 | 91 |
| 24F | 0.00223171 | 21 | 67 |
| 9V | 0.00206871 | 50 | 71 |
| 3 | 0.00002045 | 201 | 95 |
| 8 | 0.00192825 | 33 | 23 |
| 4 | 0.00192411 | 62 | 46 |
| 25A | 0.00164507 | 5 | 20 |
| 19A | 0.00155419 | 650 | 402 |
| 33F | 0.00134309 | 101 | 36 |
| 18B | 0.00126504 | 2 | 4 |
| 22F | 0.00103457 | 210 | 56 |
| 38 | 0.00088259 | 71 | 14 |
| 12B | 0.00081015 | 1 | 2 |
| 20 | 0.00077023 | 18 | 5 |
| 6B | 0.00075312 | 568 | 212 |
| 18F | 0.00073552 | 1 | 2 |
| 10A | 0.00007228 | 197 | 47 |
| 23F | 0.00069569 | 384 | 139 |
| 19F | 0.00064661 | 439 | 154 |
| 13 | 0.00064502 | 54 | 6 |
| 28F | 0.00006166 | 1 | 2 |
| 9N | 0.00052911 | 32 | 13 |
| 10B | 0.00051616 | 3 | 3 |
| 31 | 0.00049194 | 84 | 7 |
| 6A | 0.00043639 | 419 | 123 |
| 15A | 0.00041763 | 223 | 33 |
| 16F | 0.00039958 | 170 | 16 |
| 7C | 0.00039639 | 66 | 5 |
| 17F | 0.00038606 | 139 | 15 |
| 15B/C | 0.00035059 | 415 | 70 |
| 9A | 0.00033344 | 1 | 1 |
| 24A | 0.00031946 | 1 | 1 |
| 23B | 0.00026101 | 327 | 27 |
| 21 | 0.00020098 | 204 | 11 |
| 18A | 0.00019777 | 14 | 2 |
| 15F | 0.00001709 | 7 | 2 |
| 23A | 0.00001595 | 256 | 21 |
| 35F | 0.00015039 | 184 | 11 |
| 35B | 0.00013092 | 268 | 11 |
| 34 | 0.00010173 | 174 | 3 |
| 37 | 0.00008861 | 38 | 1 |
| 6C | 0.00008415 | 200 | 9 |
| 11A | 0.00006762 | 391 | 12 |
| 29 | 0.00006419 | 27 | 1 |

*Limited to serotypes where we observed carriage in Nigeria

Table S5: Summary of serotype data that contributed to CCR estimates (5) used in Model 3

| Country | Period | Number of serotypes | Number of swabs | Carriage isolates | IPD surveillance population | IPD isolates |
| --- | --- | --- | --- | --- | --- | --- |
| USA -Alabama | Pre-PCV | 11 | 827 | 103 | 19316 | 32 |
| USA - Atlanta | Post-PCV | 17 | 451 | 117 | 298831 | 47 |
| USA - Atlanta | Pre-PCV | 10 | 231 | 82 | 204680 | 192 |
| USA - Massachusetts | Post-PCV | 42 | 2969 | 792 | 820000 | 205 |
| USA - Navajo | Post-PCV | 39 | 6541 | 2046 | 65048 | 128 |
| Spain | Post-PCV | 23 | 209 | 186 | 228000 | 150 |
| Colombia | Post-PCV | 35 | 246 | 75 | 357200 | 83 |
| Colombia | Pre-PCV | 37 | 197 | 121 | 357200 | 339 |
| Venezuela | Pre-PCV | 12 | 1004 | 181 | 146125 | 33 |
| Czech | Pre-PCV | 27 | 425 | 153 | 478177 | 138 |
| England and Wales | Pre-PCV | 28 | 3752 | 648 | 3091000 | 461 |
| France | Post-PCV | 38 | 1212 | 160 | 842076 | 176 |
| France | Post-PCV | 47 | 1212 | 185 | 838866 | 388 |
| Papua New Guinea | Pre-PCV | 21 | 2844 | 416 | 96207 | 17 |
| Morocco | Pre-PCV | 8 | 200 | 33 | 212566 | 83 |
| Netherlands | Post-PCV | 36 | 659 | 328 | 222671 | 47 |
| Netherlands | Post-PCV | 37 | 660 | 364 | 232251 | 73 |
| Netherlands | Pre-PCV | 28 | 321 | 208 | 250924 | 100 |
| Canada | Pre-PCV | 7 | 1139 | 412 | 580507 | 69 |
| Portugal | Pre-PCV | 35 | 1170 | 730 | 2071223 | 90 |

Table S6: Model 1 predicted Incidence Rate Ratios (IRRs) and 95% credible intervals (CIs) for PCV10 impact on overall IPD in Nigeria comparing observed levels of non-vaccine serotype (NVT) replacement in carriage and full model assumptions of complete NVT replacement in carriage.

| Site | | NVT replacement (λ) | | IRR (95% CI) | |  | |  | |
| --- | --- | --- | --- | --- | --- | --- | --- | --- | --- |
|  | |  | | No cross protection | | Cross protection against 6A | | | |
| Kumbotso (rural) | | |  |  |  | |  | |  |
|  | Observed (0.4) | | | 0.50 | 0.47-0.54 | | 0.44 | | 0.40-0.50 |
|  | Hypothetical complete (1.0) | | | 0.69 | 0.62-0.79 | | 0.71 | | 0.60-0.84 |
| Pakoto (urban) | | | |  |  | |  | |  |
|  | | Observed (0.0) | | 0.38 | 0.36-0.39 | | 0.26 | | 0.25-0.28 |
|  | | Hypothetical complete (1.0) | | 0.73 | 0.65-0.83 | | 0.69 | | 0.59-0.82 |

**References**

1.	Johnson HL, Deloria-Knoll M, Levine OS, Stoszek SK, Freimanis Hance L, Reithinger R, et al. Systematic evaluation of serotypes causing invasive pneumococcal disease among children under five: the pneumococcal global serotype project. PLoS Med. 2010 Oct 5;7(10).

2.	Yildirim I, Hanage WP, Lipsitch M, Shea KM, Stevenson A, Finkelstein J, et al. Serotype specific invasive capacity and persistent reduction in invasive pneumococcal disease. Vaccine. 2010 Dec 16;29(2):283–8.

3.	Shea KM, Weycker D, Stevenson AE, Strutton DR, Pelton SI. Modeling the decline in pneumococcal acute otitis media following the introduction of pneumococcal conjugate vaccines in the US. Vaccine. 2011 Oct 1;29(45):8042–8.

4.	Kirkwood BR, Sterne JA. Comparing two proportions. Essential medical statistics. John Wiley & Sons; 2010. p. 148–64.

5.	Løchen A, Truscott JE, Croucher NJ. Analysing pneumococcal invasiveness using Bayesian models of pathogen progression rates. PLoS Comput Biol. 2022 Feb 17;18(2):e1009389.
